# Analysis of SARS-CoV-2 Genomes from Southern California Reveals Community Transmission Pathways in the Early Stage of the US COVID-19 Pandemic

**DOI:** 10.1101/2020.06.12.20129999

**Authors:** Wenjuan Zhang, John Paul Govindavari, Brian Davis, Stephanie Chen, Jong Taek Kim, Jianbo Song, Jean Lopategui, Jasmine T Plummer, Eric Vail

**Affiliations:** Molecular Pathology Laboratory, Department of Pathology and Laboratory Medicine, Cedars-Sinai Medical Center, Los Angeles, CA, USA; Center for Bioinformatics and Functional Genomics, Department of Biomedical Sciences, Cedars Sinai Medical Center, Los Angeles, CA, USA; Applied Genomics, Computation and Translational Core, Cedars Sinai Cancer Center, Los Angeles, CA, USA

**Keywords:** COVID-19, mutation, phylogenetic tree, SARS-CoV-2 genome sequencing, community transmission

## Abstract

Given the higher mortality rate and widespread phenomenon of Severe Acute Respiratory Syndrome Coronavirus 2 (SARS CoV-2) within the United States (US) population, understanding the mutational pattern of SARS CoV-2 has global implications for detection and therapy to prevent further escalation. Los Angeles has become an epicenter of the SARS-CoV-2 pandemic in the US. Efforts to contain the spread of SARS-CoV-2 require identifying its genetic and geographic variation and understanding the drivers of these differences. For the first time, we report genetic characterization of SARS-CoV-2 genome isolates in the Los Angeles population using targeted next generation sequencing (NGS). Samples collected at Cedars Sinai Medical Center were collected from patients with confirmed SARS-CoV-2 infection. We identified and diagnosed 192 patients by our in-house qPCR assay. In this population, the highest frequency variants were in known mutations in the 5’UTR, AA193 protein, RdRp and the spike glycoprotein. SARS-CoV-2 transmission within the local community was tracked by integrating mutation data with patient postal codes with two predominant community spread clusters being identified. Notably, significant viral genomic diversity was identified. Less than 10% of the Los Angeles community samples resembled published mutational profiles of SARS-CoV-2 genomes from China, while >50% of the isolates shared closely similarities to those from New York State. Based on these findings we conclude SARS-CoV-2 was likely introduced into the Los Angeles community predominantly from New York State but also via multiple other independent transmission routes including but not limited to Washington State and China.

## Introduction

With the emergence of the COVID-19 global pandemic caused by Severe Acute Respiratory Syndrome Coronavirus 2 (SARS-CoV-2)^1^ comes an urgent need to understand all aspects of this novel virus The SARS-CoV-2 genome sequences deposited in public databases are pivotal resources^2,3^ in understanding its virulence and for guiding approaches to therapeutics and vaccines. Assessing core genomic features across all global populations can be used to help comparative genomic analysis by identifying features unique to SARS-CoV-2 as well as assist in epidemiologic and public health endeavors.

SARS-CoV-2 is a coronavirus with a 29,903 bp single-stranded RNA (ss-RNA) genome ^4^ containing 14 ORFs and 27 predicted proteins^5^. Viral genome annotation can be used to assess the targeting of conserved unmutated sequence across all COVID-19 positive patients. A recent study summarized global distribution of SARS-CoV-2 into three central variants (A, B and C) which are distinguished by specific nucleic acid changes ^6^. The A type is thought to be the most ancestral form (T29095C). The B type (T8782C, C28144T) is most prominent in East Asia and remained there, without additional, acquired mutations, which is suggestive of founder effects or environmental resistance against this type. The C type (G26144T) is found mainly in Europeans. More specifically, sequence analysis from US East Coast patients appear to originate from the European population (C type). Whereas the US West Coast population of patients, primarily from the Seattle and Northern California area, share resemblance to A and C type^6^. While this published study is disputed, it does support the current belief that different SARS-CoV-2 genome isolates from China have seeded geographical distributions of COVID-19 throughout the US. The global consortium, NextStrain^3^ and other sequencing studies further validate the transmission of SARS-CoV-2 originated from China and disseminated to seed the US West Coast population and Europe. While Seattle recorded the first observed transmission of SARS-CoV-2 from China in the US, the largest SARS-CoV-2 US epicenter to date is New York State^7^. The isolates from New York appear to be introduced from Europe. Populations along the US West Coast, Washington State and Northern California appear to have originated directly from China. Within Northern California, another early introduction appears to occur from Washington State.

Los Angeles is the largest city on the US West Coast and the 2^nd^ major city to take precautionary measures to restrict their population to their homes as fatalities emerged early March 2020. Cedars Sinai Medical Center (CSMC) serves more than 1 million people and is the largest health service center west of the Mississippi. An in-house SARS-CoV-2 RT-qPCR diagnostic test was adopted March 21, 2020 allowing our clinical laboratory to rapidly screen and identify COVID-19 positive patients. After transmission from China, our timeline for SARS-CoV-2 testing follows other reported introductions into different global populations^8–12^. To date, the sole Los Angeles deposited SARS-CoV-2 genome is not linked to a particular mode of introduction by Nextstrain^3^. Based on these cumulative findings, we hypothesize that the local Los Angeles community was likely disseminated from a US West Coast SARS-CoV-2 strain which was directly transmitted from China. In an effort to further understand this evolving virus, we sought to perform next generation sequencing (NGS) analysis on COVID-19 positive patients. We conducted phylogenetic analyses on this unique West Coast population to identify local community spread within the greater Los Angeles area. A broader geographical distribution comparison of these early Southern California with New York State, Washington State and China isolates was conducted to ascertain possible early transmission pathways of SARS-CoV-2 dissemination into Los Angeles. Here for the first time, we report the trends for potential sources of SARS-CoV-2 introduction into the Los Angeles community.

## Methods and Materials

### Diagnostics and Sample preparation

Clinical specimens were collected by nasopharyngeal swabs from patients presenting with COVID-19 like symptoms. Total nucleic acid was extracted using the QIAamp Viral RNA Mini Kit on the QIAcube Connect (Qiagen, Germantown, USA). All patients were first assessed by CSMC developed RT-qPCR diagnostic test for SARS-CoV-2 viral RNA. Specifically, the nucleic acid was screened for the presence of SARS-CoV-2 using real-time single-plex RT-qPCR for the SARS-CoV-2 Nsp3 gene. All samples were diagnostically COVID-19 positive with amplification of the targeted region crossing the threshold before 40 cycles. In total, 189 COVID-19 positive samples were used for parallel NGS analysis.

### Targeted NGS and phylogenetic analyses

All samples were quantified by Qubit and 100 ng of total RNA was processed for 1^st^ strand and 2^nd^ strand cDNA synthesis using NEBNext Ultra II Directional RNA Library Prep Kit modular workflow (New England Biolabs, Boston, USA) according to the manufacturers’ recommendations. Target enrichment of 200 ng cDNA was performed using the Nextera Flex library preparation combined with the Illumina viral respiratory targeting panel and DNA UD indices (Illumina, San Diego, USA). After enrichment, all samples were pooled and loaded on a NovaSeq Illumina platform for paired end (150bp) sequencing All samples with greater than 50% of the genome covered with >5 depth were retained for downstream analysis (144 isolates). Duplicated reads are labelled with Picard (http://broadinstitute.github.io/picard/) and BCFtools^14^ was used to generate consensus sequence.

Global background sequences were downloaded from GISAID EpiCoV database as of May 18, 2020^2^, and only complete sequences were included, totally 3554 SARS-CoV-2 genomes. Multiple sample alignment (MSA) was performed with MAFFT (v7.464)^15^ and phylogenetic tree was generated with FastTree (v2.1)^16^ implementing generalized time-reversible (GTR) model. The final inferred tree was time scaled with TreeTime^17^.

## Results

### Sequenced SARS-CoV-2 cases from Cedars-Sinai Medical Center

We sequenced 192 specimens that are tested positive for SARS-CoV-2 using the Illumina Respiratory Virus Targeted Panel. These specimens were collected between March 22nd to April 15th including 82 females (42.7%) and 110 (57.3%) males with median age of 59.5 yrs. The overall distribution is shown in Figure 1. Of those 192 cases, 21 were deceased (10.9%), 122 were admitted and subsequently discharged (63.5%), 11 were admitted and currently hospitalized for treatment (5.7%), and 38 were outpatients that were not hospitalized for COVID-19 (19.8%). The pool of 192 SARS-CoV-2 positive samples obtained 2,222,425,974 reads in raw data. A minimum of ∼1M reads per sample was generated and mapped to the SARS-CoV-2 reference. A total of 1,737,684,077 reads (78% of total) were mapped to SARS-CoV-2 reference genome (NC_045512.2) obtained from NCBI GenBank using BWA-MEM (0.7.17-r1188)^13^ with default parameters. Mapping ratio varies between 0.3% to 98.98% which positively correlated (R_2_=0.42) with Ct value obtained from RT-qPCR. Overall, low mapping ratios that fell below 50% genome coverage correlated to samples which Ct value (>30 cycles) in the RT-qPCR diagnostic test.

**Figure 1.**
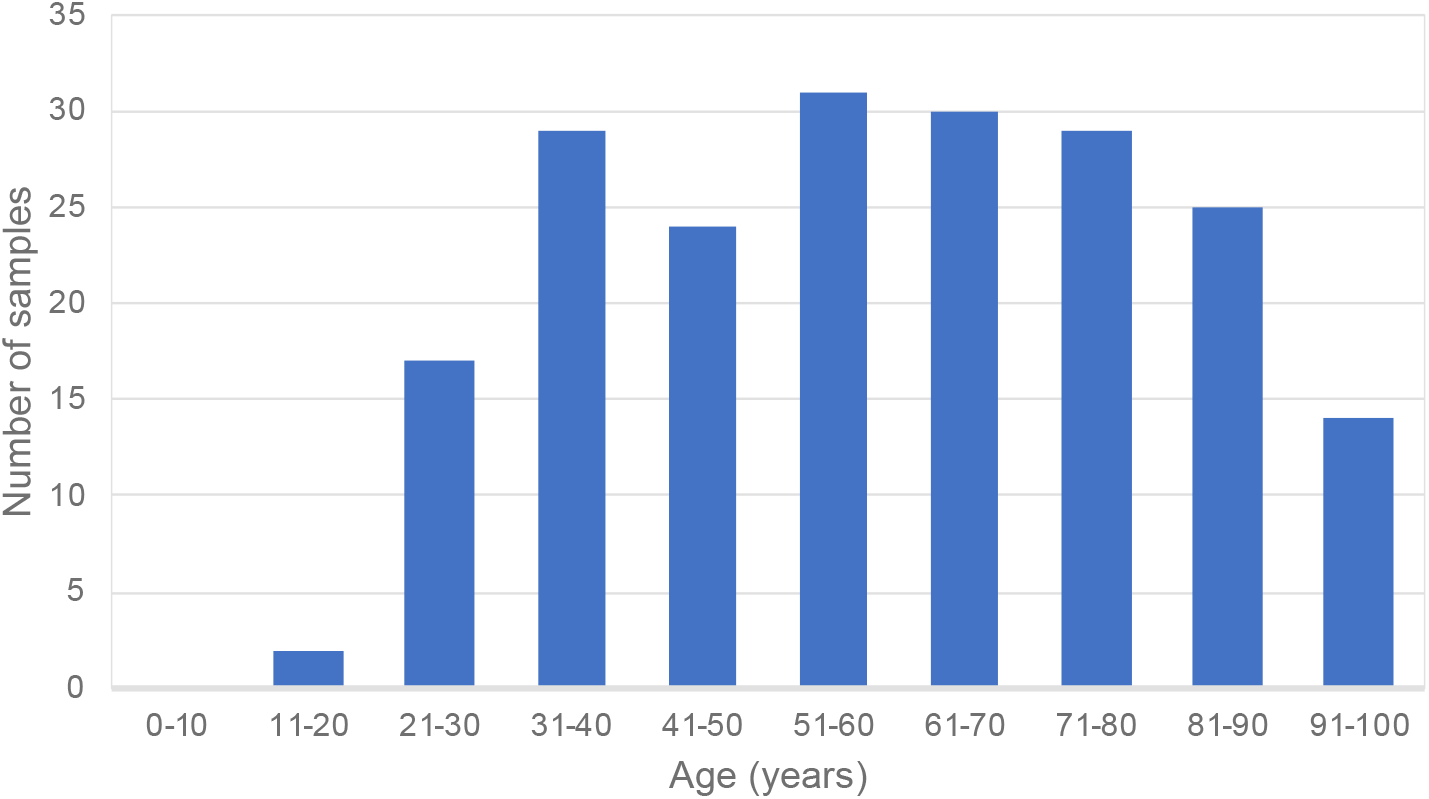
Histogram demonstrating the age demographic of SARS-CoV-2 positive patients sampled from Cedars Sinai Medical Center in Los Angeles, USA.

### SNP profile consistent with SARS-CoV-2 signatures from Europe and US isolate signature

Whole-genome comparison of the CSMC samples revealed >99.8% identity with the SARS-CoV-2 reference genome (NC_045512.2). Mutational analyses of this sample set against the reference genome revealed a total of 518 mutated sites detected across the length of the SARS-CoV-2 genome (Figure 2). 84% of the variants were private and five variants were found in more than half of all samples sequenced (Table 1). In total, we identified 82 sites that were mutated in more than two isolates in this cohort. These CSMC isolates contained on average 5.1 mutations per sample. The top 20 mutated sites, their predicted alterations and frequencies are summarized in Table 1 and Supplemental Figure 1.

**Table 1.** Top 20 mutations of the SARS-CoV-2 genome discovered in the samples collected from Cedars Sinai Medical Center. Mutation site is depicted in base pairs (bp) along the genome sequence of SARS-CoV-2 and predicted the amino acid (aa) alteration of the corresponding protein. M-ain monophyletic clades were labelled based on nucleotide substitutions.

| Pos | Ref | Alt | Gene | Protein | Amino Acid Substitutions | Clade |
| --- | --- | --- | --- | --- | --- | --- |
| 241 | C | T | 5'UTR | 5'UTR | NA |  |
| 313 | C | T | ORF1ab | nsp1 | synonymous |  |
| 379 | C | A |  | nsp1 | synonymous |  |
| 1059 | C | T |  | nsp2 | T>I |  |
| 3037 | C | T |  | nsp3 | synonymous |  |
| 8782 | C | T |  | nsp4 | synonymous |  |
| 11083 | G | T |  | nsp6 | L>F |  |
| 13575 | T | C |  | RdRp | synonymous |  |
| 14408 | C | T |  | nsp12 | synonymous | A2a |
| 17747 | C | T |  | nsp13 | P>L |  |
| 17858 | A | G |  | helicase | Y>C |  |
| 18060 | C | T |  | 3'-to-5'exonuclease | synonymous | B1 |
| 18877 | C | T |  | nsp14 | synonymous |  |
| 23403 | A | G | S | spike glycoprotein | D>G | A2 |
| 25466 | C | T | ORF3a | ORF3a protein | P>L |  |
| 25563 | G | T | ORF3a | ORF3a protein | Q>H |  |
| 28144 | T | C | ORF8 | ORF8 protein | L>S | B |
| 28881 | G | A | ORF9/ N | nucleocapsid phosphoprotein | R>K |  |
| 28882 | G | A,T | ORF9/ N | nucleocapsid phosphoprotein | R>K* |  |
| 28883 | G | C | ORF9/ N | nucleocapsid phosphoprotein | G>R |  |

**Figure 2.**
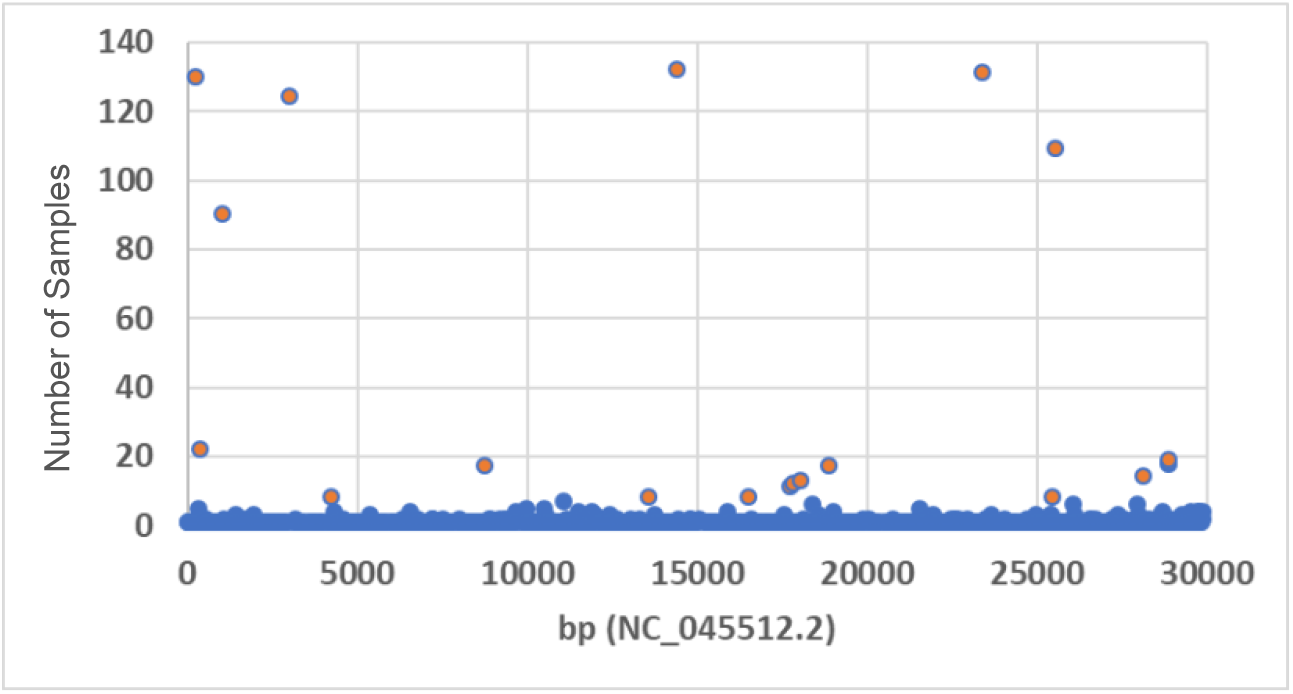
Frequency dot plot demonstrating the number of samples with mutation sites along the length (bp, base pair) of the SARS-CoV-2 genome (NC_045512.2). Red circles depict top 20 mutated sites seen in the Cedars Sinai Medical Center samples.

From our most observed mutation sites, four variants have been previously reported including in the 5’-UTR (C241T), along with C3037T, C14408T, A23403G^18^ and we found 125 (65%) samples with all four variants presented in the genome. While C3037T causes a synonymous mutation in nsp3 (F105F), C14408T and A23403G resulted in amino acid changes in RNA primase (nsp12, P323L), and S protein (D614G), respectively. Mutations at G25563T (ORF3a) and C1059T (nsp2) have been reported to be co-expressed. Furthermore, Type A (East Asia) and C (European) mutations were not among the highest mutations defining the CSMC isolates. Type B is defined by two mutations but only the C28144T mutation was frequently observed in our population. These results are inconsistent with published findings of a Chinese isolate being the main transmission source for the Los Angeles community spread.

### Phylogenetic analysis reveals multiple introductions to Los Angeles county from diverse origins

After quality assurance of sequencing reads, 144 samples which have greater than 50% of the genome covered with greater than 5X genome depth were retained for phylogenetic analysis. We performed phylogenetic analysis of the 144 CSMC samples to identify which genomes most closely resemble each other (Figure 3). These CSMC SARS-CoV-2 genomes were time scaled with specimen collection date (Supplemental Figure 2). By labeling the top six mutated sites along the phylogenetic tree, (Figure 4) we observed a minimum of two groups containing distinct variant signatures. Within these groups, the presence of all six variants exist in genome isolates from the bottom subclade of the tree (Figure 4A-F). Interestingly, a subset of the four variants that track together, as previously described, fall into these two main clusters (Figure 4A, C, D, E). While these variants tightly segregate into two main clusters of the tree, they do not appear to track with sample date collection (Supplemental Figure 2). The genomic diversity in our population was an early introduction into the community and remained through the timing of collections.

**Figure 3.**
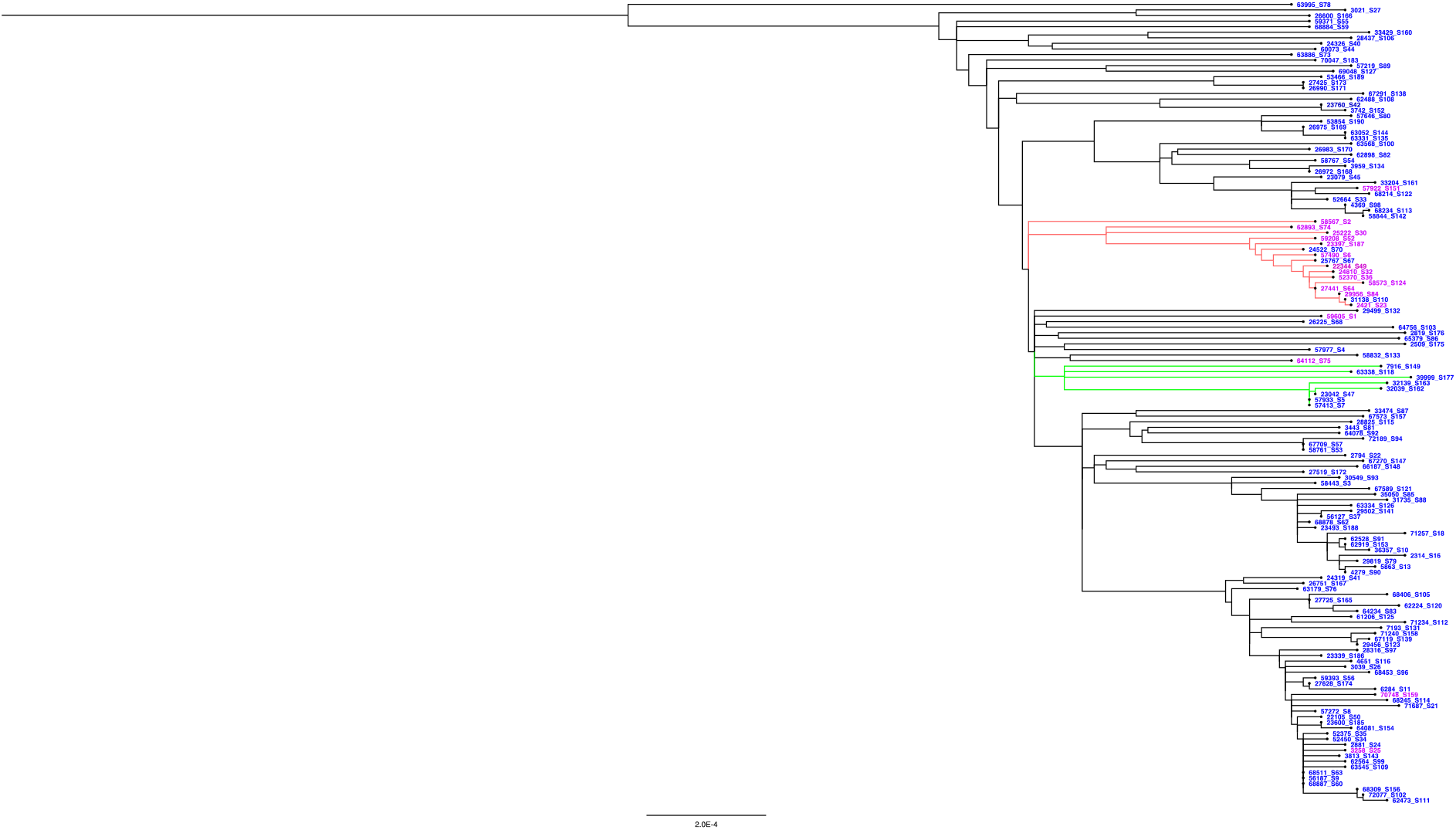
Phylogenetic tree of 144 SARS-CoV-2 genomes sampled from Cedars Sinai Medical Center patients in Los Angeles, USA, collected from March 22nd to April 15^th^ 2020.

**Figure 4.**
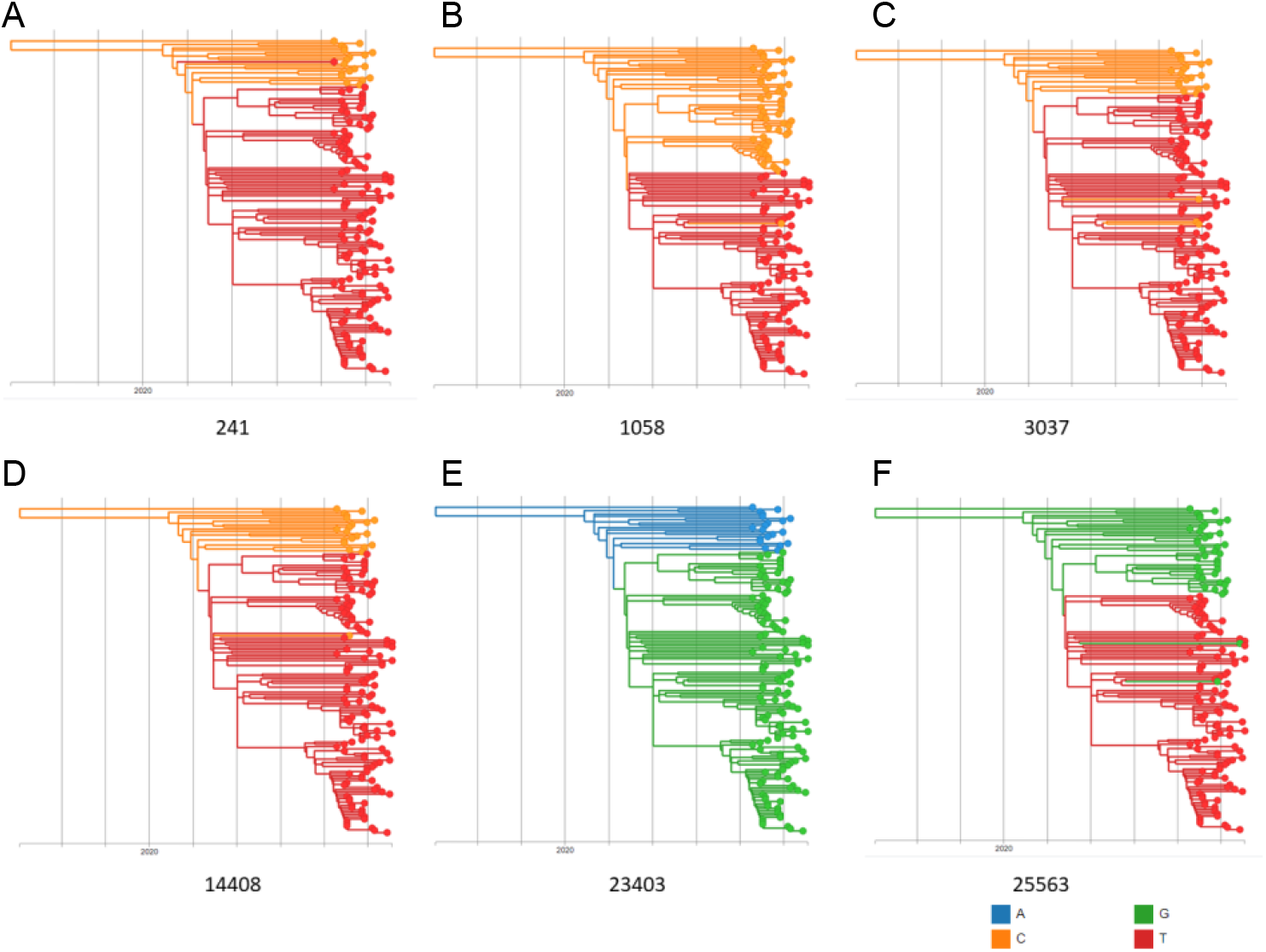
Phylogenetic tree of the six most frequently mutated sites observed in the SARS-CoV-2 genomes of Cedar Sinai Medical Center samples. Each nucleotide has a corresponding color code (A-blue, G-green, C-orange, T-red), Each histogram depicts two main clusters seen throughout all six variant changes.

### CSMC phylogenetic tree traces back community transmission in the early stage of the COVID-19 epidemic

From our local phylogenetic tree analysis, 17 patients which represents >10% of our sample population were identified in one cluster (red) (Figure 3). Further meta-data analysis revealed, these patents lived in the same or adjacent postal code and were all members of the same religious denomination. An additional community transmission cluster was observed with a tightly associated cluster containing eight patients (green) (Figure 3) while the viral genome of these patients share variant C1887T exclusively (Supplemental Figure 3). We did not observe other obvious connections within samples outside of those two clusters.

### Joint phylogenetic analysis revealed genetic similarity with NYC isolates

To properly address the route of transmission and the distribution of the Los Angeles population in relation to the global distribution of the SARS-CoV-2, we combined the CSMC samples with genomes isolated from possible places of introduction: New York City, Washington State and China. All complete SARS-CoV-2 genomes from these locations deposited in GISAID (https://www.gisaid.org/) until May 18th, 2020 were used for the subsequent analyses (Supplemental Figure 4). The phylogenetic tree reveals several clusters (A-G) which are labeled based on the geographic distribution of the SARS-CoV-2 genomes (Figure 5). From this joint phylogenetic analysis, our local Los Angeles samples were distributed throughout all clusters of the phylogenetic tree with the exception of F. The distribution of CSMC samples amongst these three geographically distributed isolates is indicative of multiple independent introduction events into the local Los Angeles community. A large percentage (67%) of the CSMC SARS-CoV-2 genomes cluster within clusters A, E and G which contain predominantly New York City isolates. In fact, a scattering of CSMC isolates are found in all three New York clusters. Similarly, there are two main clusters (B and D) from China in which CSMC samples exist in both clusters, with 7% of the total CSMC samples closely resemble isolates from China. The remaining isolates (26%) lie within cluster D defined by Washington State deposited SARS-CoV-2 genomes. Interestingly the only cluster in which SARS-CoV-2 genomes from CSMC are excluded is one of the two Washington State clusters (F).

**Figure 5.**
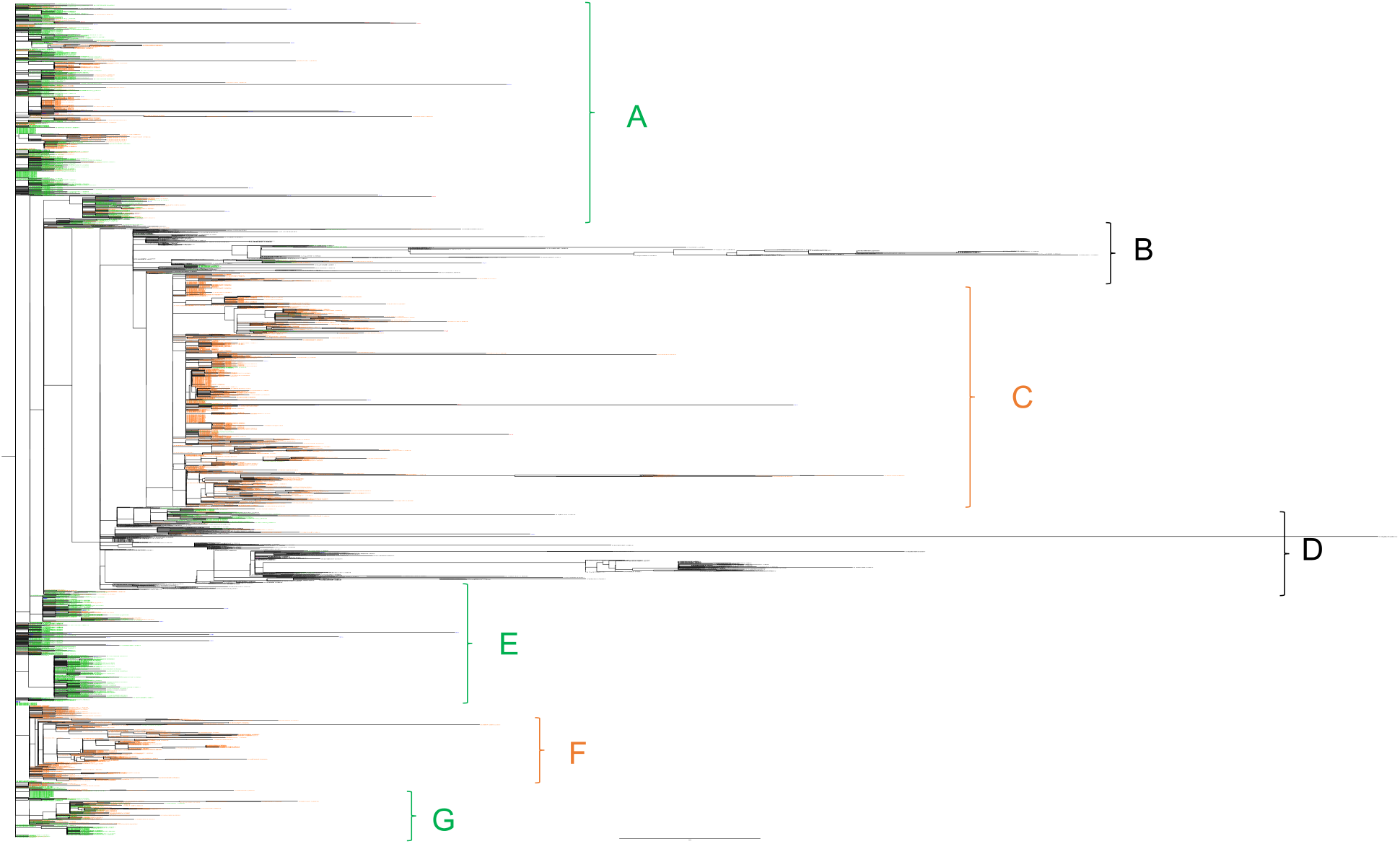
Phylogenetic tree of 3354 SARS-CoV-2 genomes. This tree is drawn to compare patient genomes Washington State(orange), New York State (green) and China black) to those sequenced from Cedars Sinai Medical Center patients in Los Angeles, USA (blue).

## Discussion

We present for the first time a comprehensive study of sample population from one of the COVID-19 epicenters in the US. A caveat to our sample collection is that the emergency department admissions are less frequent for younger patients and biased to patients greater than 18 years old. Despite this, the average age of CSMC patients was ∼60 years old, which is consistent with other observations that older adults are susceptible to COVID19^10^. Patient samples by diagnostic RT-qPCR that had lower cycle numbers e.g. higher viral copies generally had poorer outcomes. Moreover, patients with higher viral loads detected by RT-qPCR also correlated to a higher percent SARS-CoV-2 genome coverage by sequencing. From a technical perspective, 48 of 192 patients which had poorer quality sequencing coverage (<50%) and also were diagnostically positive by RT-qPCR at >30 cycles. The remaining144 patients passed all quality control metrics for sequencing and alignment generally <25 cycles of RT-qPCR^19^. Hence, when using NGS approaches for diagnostic purposes, a potential caveat is that genome sequencing favors those patients with higher viral titers and may not capture those who have low viral copy numbers.

The local phylogenetic tree demonstrated two large clusters which were mainly defined by six high frequency mutations. Phylogenetic analysis of these samples by collection date, reveals the main variants which define these two large clusters were observed throughout March and April and were present in the community prior to the collection start date. This study presents a snapshot of the molecular characteristics of SARS-COV-2 early transmission into the Los Angeles area. Based on the window of our collection dates, it was likely not sufficient time to observe new dissemination into this population. Despite our local phylogenetic tree showing high genomic diversity, we also observed tight clustering patterns within including a tight cluster (green) of eight patients in which their genomes shared one variant in common This finding directly highlights the precision of contact tracing directly through SARS-CoV-2 genome isolation. Moreover, another unique cluster in the local phylogenetic tree, found 17 patients who were identified within the same postal code. This postal code is only 2.37 square miles and highly populated (36,885 people). This selected cluster represents spread within a constrained geographic area all within members of a religious community. It has been noted previously how religious communities are of particular risk in a pandemic such as this due to large communal events such as services, weddings and funerals. Moving forward community leaders should be aware of the unique risks posed to their congregations and should plan accordingly. Moreover, the remaining samples live across many postal codes, providing further evidence of community transmission across the larger metropolitan area. Global initiatives to track SARS-CoV-2 throughout the world have proven fruitful in monitoring disease incidence, severity and worldwide spread^6,20–24^. In this study by examining a cohort within a SARS-CoV-2 US epicenter, Los Angeles, we lay the foundation for further studies into the use of SARS-CoV-2 sequencing to monitor local community spread.

Although we have a limited sample number (192 COVID-19 positive patients), the integration of our local US West Coast SARS-CoV-2 genomes into Washington State, New York State and China, provided helpful insight into determining the introduction of SARS-CoV-2 to this Los Angeles community. Consistent with other studies, the combination of the four variants(C241T, C3037T, C14408T, A23403G) co-evolving and tracking together has been seen in other populations tracking in European isolates^18^. From our variant analysis, two of our highly mutated sites, G25563T (ORF3a) and C1059T (nsp2) have been reported exclusively in US isolated sequences collected since March 2020 ^25^, a timeline which corresponds to this study’s sample collection date. Moreover, these variants were found to be closely associated within a cluster containing mainly SARS-CoV-2 genomes from New York State, suggesting that these genomes were introduced from a strain that emerged from the US East Coast population. From the variants found in our samples, the four variants: 5’-UTR (241C>T), along with 3037C>T, 14408C>T, 23403A>G reproduce other studies in which these mutations co-evolve^18^. Given such a high proportion of our patients have all 4 mutations indicates the seeding of our population by a strain originating in Europe.

This finding is further validated in: 1) our local phylogenetic tree which disseminates into 2 main clusters and 2) our global tree in which our population closely resembles SARS-CoV-2 genomes geographically distributed with the majority from New York City, followed by a large fraction from Washington State, together identifying possible routes for dissemination to Southern California. Given Seattle was the 1st documented US appearance of SARS-CoV-2 the introduction of the virus from Washington State^7^ is consistent with our tree and the timing of our data sampling, consistent with our hypothesis. However, despite our earlier predictions an even larger portion of our sample population had a high resemblance to genomes from New York State. With New York City being the largest epicenter of the SARS-CoV-2^7,26,27^ and the appearance of our samples within 3 separate clusters of New York isolates, SARS-CoV-2 likely disseminated from multiple introductions from New York State. Furthermore, the CSMC population also clustered tightly within a large Washington State cluster likely disseminated to Southern California, appearing as a major cluster in our local population. Although we restricted our analyses to these three geographical origins, given we found a high genomic diversity amongst the CSMC SARS-CoV-2 isolates, COVID-19 large impact on the Los Angeles community likely originated from independent disseminations of the virus from multiple geographical routes.

## Data Availability

All data will be deposited in the publicly available database GISAID www.gisaid.org

## Acknowledgements

The authors would like to thank all those that are helping in the fight against SARS-CoV-2 especially the research community who has kindly shared genomic data, publicly available for download from www.gisaid.org.

## Author Contributions

WZ, JP and EV conceived of the project and experimental design. JP, BS, SC executed the experiment and generated the sequencing data. WZ conceived and executed all data analyses. WZ, JP and EV contributed to the writing of the manuscript. JL, JK, JS and JPG gave helpful insight and suggestions from conception through to execution and writing.

## Funding

This project was funded by an internal grant to Eric Vail generously provided by the Department of Pathology and Laboratory Medicine, Cedars Sinai Medical Center.

## Conflict of Interest

The authors declare no conflicts of interest.

## Notes

### Competing Interest Statement

The authors have declared no competing interest.

### Author Declarations

Cedars-Sinai's Institutional Review Board (IRB) reviewed and approved all research described herein.

